# Leishmaniasis on YouTube: a critical appraisal of the quality, reliability, and transparency of educational content

**DOI:** 10.64898/2026.06.18.26355931

**Authors:** Kaweesi Calvin Nantalaga, Nantege Alice

## Abstract

**Background:** Leishmaniasis is a neglected tropical disease of significant global public health importance, for which accurate information is essential to support prevention and early care-seeking, particularly in endemic, resource-limited settings. YouTube is a widely used source of health information, but the quality and reliability of leishmaniasis-related content have not been evaluated. We aimed to assess the quality, reliability, and transparency of English-language YouTube videos on leishmaniasis.

**Methods:** We conducted a cross-sectional analysis of YouTube videos retrieved via the YouTube Data API on 15 June 2026 using the terms “leishmaniasis,” “cutaneous leishmaniasis,” and “visceral leishmaniasis.” After applying eligibility criteria and screening the 150 most-viewed eligible videos, 48 videos were included. Two reviewers independently assessed each video using the modified DISCERN (mDISCERN) tool, the Global Quality Score (GQS), and the JAMA benchmark criteria, with disagreements resolved by consensus. Inter-rater agreement was assessed using the intraclass correlation coefficient (ICC), and associations were examined using Spearman’s rank correlation.

**Results:** Of 402 videos retrieved, 48 met the inclusion criteria. The median GQS was 3.00 (IQR 2.00–4.00) and median mDISCERN was 3.00 (IQR 2.38–4.50), indicating moderate quality and reliability, while the median JAMA score was 2.00 (IQR 1.00–2.00), reflecting limited transparency; no video met all four JAMA criteria. The overwhelming majority of videos (47/48, 97.9%) were of professional or institutional origin. Inter-rater agreement was good to excellent (ICC 0.883 for GQS, 0.896 for mDISCERN, 1.000 for JAMA). The instruments were strongly inter-correlated (mDISCERN–GQS ρ = 0.841, p < 0.001). Quality scores did not correlate positively with views, likes, or video duration; comments correlated weakly and negatively with mDISCERN (ρ = −0.337, p = 0.031) and JAMA (ρ = −0.381, p = 0.014).

**Conclusions:** YouTube videos on leishmaniasis are of moderate quality and reliability but limited transparency, and are produced almost exclusively by professional sources. Video popularity, length, and age were not indicators of quality. There is a need for experts and institutions to produce clearly authored, well-sourced, and transparent educational content on this neglected tropical disease.

**Author summary:** Leishmaniasis is a neglected tropical disease caused by parasites and spread by sandfly bites, and accurate public information is important for its prevention and early treatment, especially in the poorer regions where it is most common. Many people turn to YouTube for health information, but the quality of leishmaniasis videos had never been studied. We analysed 48 English-language YouTube videos on leishmaniasis using three established scoring tools assessing quality, reliability, and transparency. We found that the videos were of moderate quality and reliability but were often not transparent about who made them, what sources they used, or when they were produced; no video met all four transparency criteria. Almost all the videos came from health professionals or institutions rather than ordinary users, which differs from many other health topics on YouTube. Importantly, a video’s popularity, length, or age did not indicate how good it was. Our findings suggest that experts and health institutions should produce not only more, but clearer and better-sourced, educational content on leishmaniasis to help people find trustworthy information online.

## Introduction

Leishmaniasis is a vector-borne neglected tropical disease (NTD) caused by protozoan parasites of the genus Leishmania and transmitted through the bite of infected female phlebotomine sandflies [1]. It manifests in three principal clinical forms: cutaneous leishmaniasis, the most common, which produces skin lesions and lasting scarring; mucocutaneous leishmaniasis, which destroys the mucous membranes of the nose, mouth, and throat; and visceral leishmaniasis (also known as kala-azar), the most severe form, which is fatal in the majority of cases if left untreated [1]. The disease is endemic across large parts of Africa, the Americas, the Mediterranean basin, the Middle East, and Asia, with an estimated 700,000 to one million new cases each year, and disproportionately affects poor and marginalised populations, reflecting its status as a neglected disease of global public health importance [1,2].

Effective control of leishmaniasis depends not only on diagnosis and treatment but also on accurate public and professional understanding of the disease, particularly in endemic and resource-limited settings where conventional health-education resources may be scarce. In this context, digital platforms have become increasingly important sources of health information. YouTube, one of the most widely used video-sharing platforms worldwide, offers freely accessible audiovisual content that can reach large and geographically dispersed audiences, making it an attractive medium for disseminating health information. However, unlike peer-reviewed literature, content uploaded to YouTube is not subject to formal review, and its accuracy and reliability vary considerably. Viewers are therefore left to judge the trustworthiness of what they watch, often influenced by their own health and digital literacy.

A growing body of research has appraised the quality of health-related YouTube content across numerous conditions, frequently using validated instruments such as the modified DISCERN tool, the Global Quality Scale, and the JAMA benchmark criteria [3–5]. These studies have repeatedly shown that video quality is uneven and that popularity is a poor proxy for accuracy [6–9]. For neglected tropical diseases specifically, such appraisals remain scarce, and for leishmaniasis, despite its substantial global burden and its reliance on public awareness for prevention and early care-seeking, no systematic evaluation of the quality and reliability of YouTube content has been conducted. We therefore aimed to assess the quality, reliability, and transparency of English-language YouTube videos on leishmaniasis, and to examine whether these attributes are associated with video characteristics and viewer engagement.

## Materials and methods

### Study design and search strategy

We conducted a cross-sectional analysis of YouTube videos providing information on leishmaniasis. Data retrieval was performed on 15 June 2026 using the YouTube Data API (v3) accessed through a script developed in R (version 4.5.3). Three search terms were queried: “leishmaniasis,” “cutaneous leishmaniasis,” and “visceral leishmaniasis.” For each term, up to 200 video records were retrieved through automated, paginated API queries (50 results per request), with results ranked by YouTube’s default relevance setting. For every video, the script extracted the video identifier, title, channel, upload date, view count, like count, comment count, and duration. After removal of duplicates across search terms, 402 unique videos were identified for screening.

### Eligibility criteria

Videos were eligible for inclusion if they (i) contained information related to leishmaniasis, (ii) were freely accessible, and (iii) were in the English language. Videos were excluded if they were shorter than one minute or longer than one hour, had fewer than 100 views, lacked audio, were advertisements or fundraising campaigns, were unrelated to leishmaniasis, were duplicates, or consisted of pronunciation guidance only. Duration and view-count thresholds were applied automatically within the R script; the remaining criteria were assessed manually. These thresholds are consistent with prior YouTube quality appraisals, reflecting that very short videos provide insufficient educational content, very long videos promote viewer disengagement, and videos with minimal views fall outside the range typically accessed by users [6].

### Screening

Of the 402 retrieved videos, 120 were automatically excluded for failing the duration or view-count thresholds, leaving 282 eligible videos. Given the volume of results and consistent with established YouTube analyses, the 150 most-viewed eligible videos were selected for detailed manual screening, reflecting the observation that users seldom view beyond the most popular results. These 150 videos were screened against the full eligibility criteria, and 102 were excluded (98 not in English, 3 lacking audio, and 1 duplicate), yielding a final sample of 48 videos for assessment (Fig 1).

**Fig 1.**
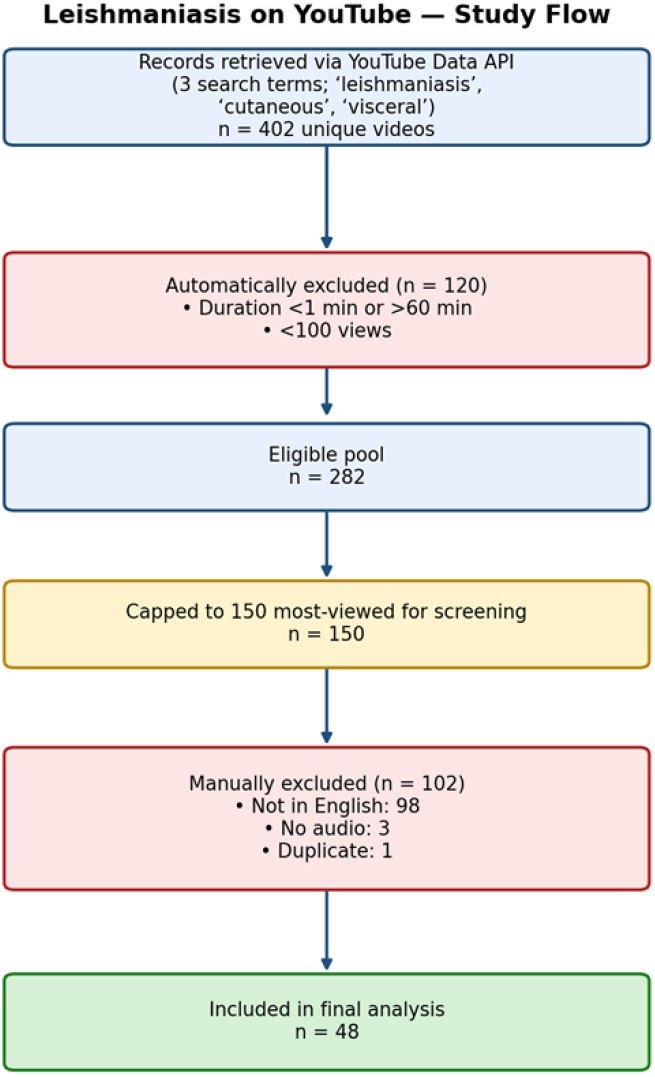
Study flow diagram showing video retrieval, screening, and inclusion.

### Quality and reliability assessment

Each included video was independently assessed by two reviewers using three validated instruments. Reliability was evaluated using the 5-item modified DISCERN (mDISCERN) tool, in which each item is scored 1 (yes) or 0 (no), giving a total of 0–5, with higher scores indicating greater reliability [3]. Overall quality was assessed using the 5-point Global Quality Score (GQS), where scores of 1–2 denote low quality, 3 moderate quality, and 4–5 high quality [4]. Transparency and credibility were assessed using the four JAMA benchmark criteria (authorship, attribution, disclosure, and currency), each scored as present (1) or absent (0), for a total of 0–4 [5]. Source-related items and attribution were scored as present if the relevant information appeared within the video or its description. Each video was further classified by upload source as professional (uploaded by a healthcare institution, educational body, medical organisation, or an identifiable health professional) or consumer-generated (uploaded by an individual without an identifiable professional affiliation).

The two reviewers scored all 48 videos independently and blinded to each other’s ratings. Disagreements were resolved by consensus to derive the final scores used in the analysis.

### Statistical analysis

Analyses were performed in R (version 4.5.3). Continuous variables were summarised as medians and interquartile ranges (IQR). Inter-rater agreement between the two reviewers was assessed using the intraclass correlation coefficient (ICC; two-way random-effects model, absolute agreement), with values below 0.50 indicating poor, 0.50–0.75 moderate, 0.75–0.90 good, and above 0.90 excellent reliability. Spearman’s rank correlation was used to examine associations between quality scores (GQS, mDISCERN, JAMA) and video metrics (views, likes, comments, duration, and time since upload), as well as between the scoring instruments. Videos with disabled likes or comments were excluded pairwise from the relevant correlations. Statistical significance was set at p < 0.05.

### Ethics statement

This study analysed publicly available, non-identifiable content on a public platform and did not involve human participants, personal data, or interaction with individuals. Ethical approval was therefore not required.

## Results

### Screening and video characteristics

A total of 402 unique videos were retrieved across the three search terms. After automatic exclusion of 120 videos failing the duration or view-count thresholds, 282 videos remained eligible, of which the 150 most-viewed were screened in detail. A further 102 videos were excluded (98 not in English, 3 without audio, and 1 duplicate), yielding 48 videos for final analysis (Fig 1).

The 48 included videos had a median of 10,716 views (IQR 3,756–35,842), 153 likes (IQR 48– 647), and 8 comments (IQR 3–31). The median video duration was 7.3 minutes (IQR 2.7–16.3), and the median time since upload was 1,889 days (IQR 1,252–2,979). Likes were disabled or hidden for 3 videos and comments for 7. Notably, 47 of the 48 videos (97.9%) were classified as professional or institutional in origin, with only a single consumer-generated video meeting the inclusion criteria; the planned comparison of quality by upload source was therefore not statistically meaningful and is not reported (Table 1).

**Table 1.** Characteristics of the 48 included leishmaniasis videos. Variable Median.

| Variable | Median (IQR) or n (%) |
| --- | --- |
| Views | 10,716 (3,756–35,842) |
| Likes (n = 45) | 153 (48–647) |
| Comments (n = 41) | 8 (3–31) |
| Duration, minutes | 7.3 (2.7–16.3) |
| Days since upload | 1,889 (1,252–2,979) |
| Global Quality Score (1–5) | 3.00 (2.00–4.00) |
| mDISCERN (0–5) | 3.00 (2.38–4.50) |
| JAMA benchmark (0–4) | 2.00 (1.00–2.00) |
| Source: professional | 47 (97.9%) |
| Source: consumer | 1 (2.1%) |

### Quality and reliability

The overall quality of the videos was moderate. The median GQS was 3.00 (IQR 2.00–4.00) and the median mDISCERN score was 3.00 (IQR 2.38–4.50), indicating moderate quality and reliability, while the median JAMA score was 2.00 (IQR 1.00–2.00), reflecting limited transparency; no video met all four JAMA criteria. The distribution of scores across the three instruments is shown in Fig 2; GQS ratings clustered at the moderate-to-good end of the scale, whereas JAMA scores were concentrated at the lower end.

**Fig 2.**
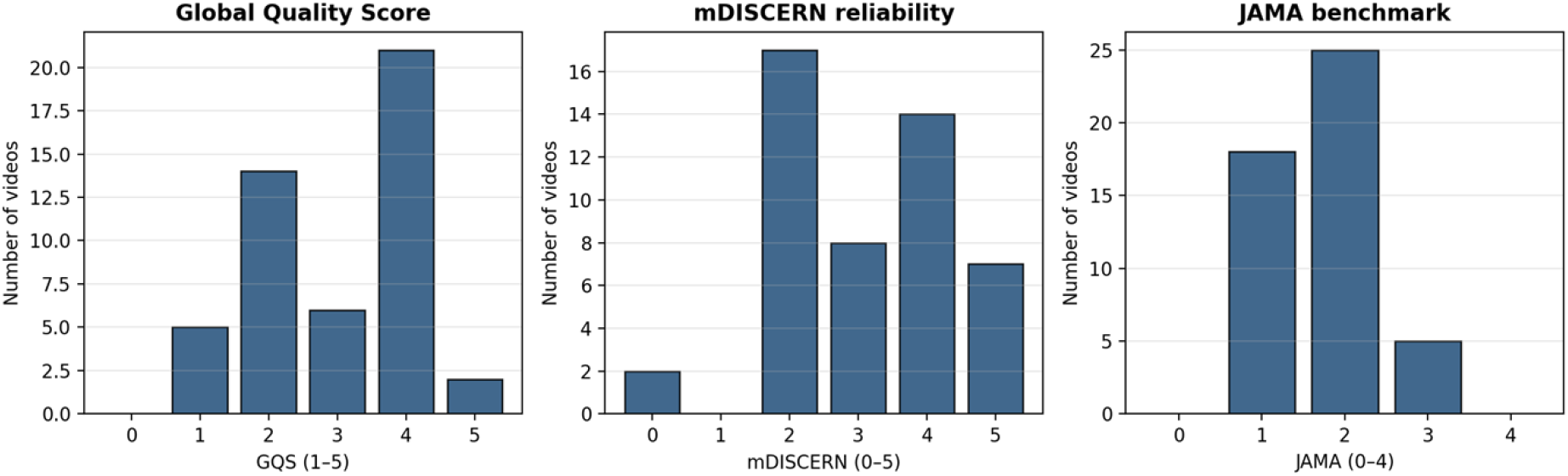
Distribution of Global Quality Score, mDISCERN, and JAMA benchmark scores across the 48 videos. Consensus (two-reviewer mean) scores are rounded to the nearest whole number for display.

### Inter-rater reliability

Agreement between the two independent reviewers was good to excellent. The ICC was 0.883 for GQS and 0.896 for mDISCERN (both good), and 1.000 for the JAMA benchmark criteria (excellent), supporting the consistency of the assessments.

### Correlation analysis

The three instruments were strongly and significantly inter-correlated: mDISCERN correlated with GQS (ρ = 0.841, p < 0.001), and the JAMA score correlated with both GQS (ρ = 0.587, p < 0.001) and mDISCERN (ρ = 0.652, p < 0.001), indicating that videos rated as higher quality also tended to be more reliable and more transparent. In contrast, quality scores were not positively associated with video popularity or other metrics. None of GQS, mDISCERN, or JAMA correlated significantly with views, likes, video duration, or time since upload (Table 2, Fig 3). The number of comments showed weak negative correlations with both mDISCERN (ρ = −0.337, p = 0.031) and the JAMA score (ρ = −0.381, p = 0.014). No significant association was observed between video duration and any quality measure.

**Table 2.** Spearman correlations between quality scores and video metrics (n = 48).

| Video metric | GQS $\rho$ (p) | mDISCERN $\rho$ (p) | JAMA $\rho$ (p) |
| --- | --- | --- | --- |
| Views | 0.193 (0.188) | -0.023 (0.875) | -0.216 (0.140) |
| Likes | 0.159 (0.297) | -0.047 (0.760) | -0.188 (0.217) |
| Comments | -0.135 (0.401) | -0.337 (0.031)* | -0.381 (0.014)* |
| Duration (min) | -0.116 (0.432) | -0.109 (0.461) | -0.085 (0.564) |
| Days since upload | -0.172 (0.242) | -0.046 (0.756) | -0.246 (0.091) |

\* $p < 0.05$ . Inter-scale: mDISCERN–GQS $\rho = 0.841$ \*; JAMA–GQS $\rho = 0.587$ \*; JAMA–mDISCERN $\rho = 0.652$ \*. Inter-rater ICC: GQS 0.883, mDISCERN 0.896, JAMA 1.000.

**Fig 3.**
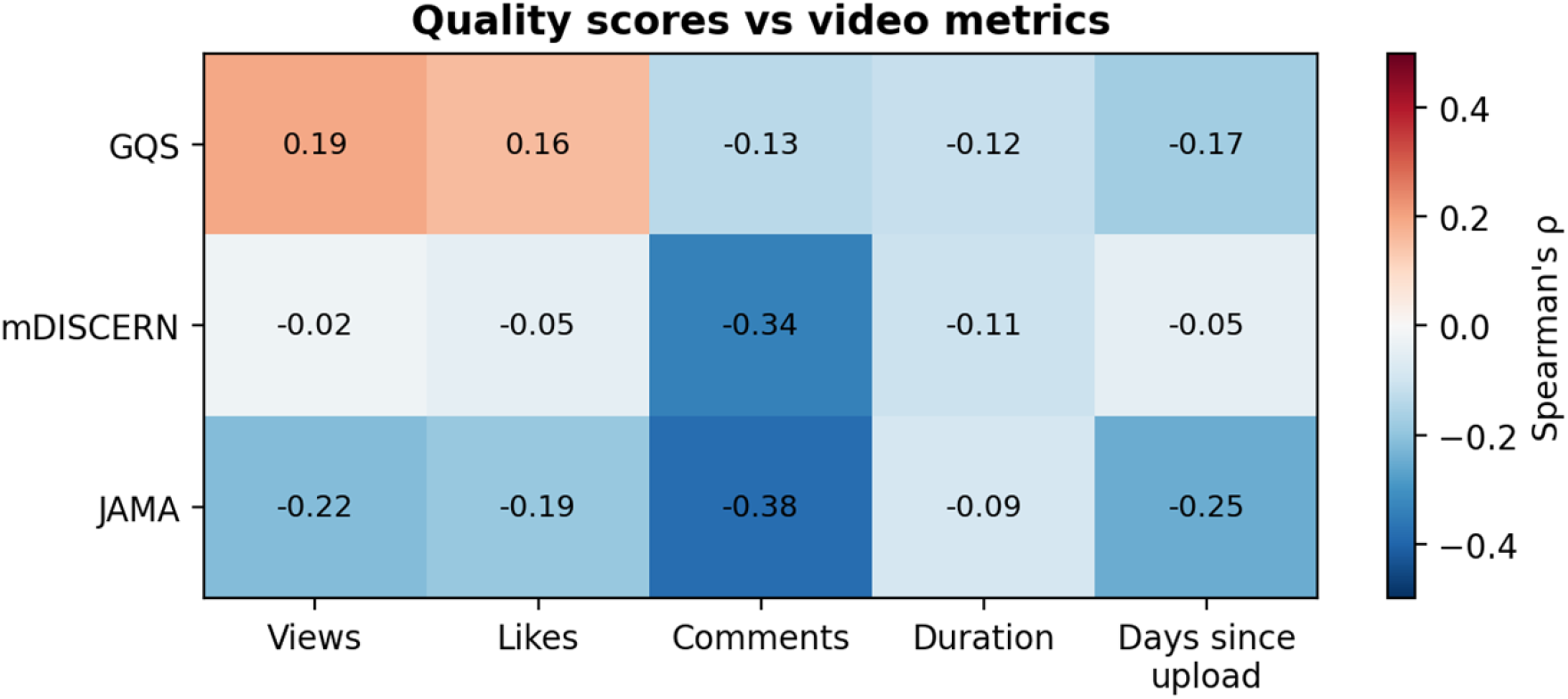
Heatmap of Spearman correlations between quality scores and video metrics.

## Discussion

To our knowledge, this is the first study to evaluate the quality, reliability, and transparency of YouTube videos on leishmaniasis. We found that English-language content on this neglected tropical disease was of moderate overall quality and moderate reliability, with notably limited transparency. The three instruments were strongly inter-correlated, inter-rater agreement was good to excellent, and, importantly, quality bore no positive relationship to a video’s popularity, length, or age.

The moderate quality we observed (median GQS and mDISCERN both 3.0) is broadly consistent with appraisals of YouTube content for other conditions, which have repeatedly described mediocre quality and reliability. Studies of envenomation and poisoning content, for example, reported comparable mid-range mean GQS and mDISCERN values and similarly modest JAMA transparency scores [7]. Our JAMA findings were particularly striking: no video met all four benchmark criteria, and the median score was 2 of 4, indicating that authorship, attribution, disclosure, and currency were frequently absent. This transparency gap mirrors that reported across the wider YouTube health literature [5,7] and is concerning, because the absence of clear authorship and sourcing makes it difficult for viewers to judge the credibility of what they watch.

A distinctive feature of our sample was its near-total dominance by professional and institutional uploaders, with 47 of 48 videos (97.9%) originating from healthcare institutions, educational bodies, or identifiable health professionals, and only a single consumer-generated video meeting the inclusion criteria. The professional dominance we observed likely reflects the clinical and unfamiliar nature of leishmaniasis: it is not a condition that lends itself to consumer commentary, and the most-viewed English-language material is produced largely by veterinary, medical-education, and institutional channels. While this is reassuring in that the available content is mostly expert-generated, the only moderate quality and poor transparency of even this professional content indicate clear room for improvement. It also meant that the planned comparison of quality by source could not be meaningfully performed, in contrast to studies of other topics where source was a significant determinant of quality [6,8].

Consistent with a large body of prior work, we found that viewer engagement metrics were not positive indicators of quality. Views, likes, and video duration showed no significant positive correlation with any quality measure, reinforcing that popularity is not a reliable proxy for accuracy [6,7]. If anything, more heavily commented videos tended to be marginally less reliable and less transparent. This has practical implications: a viewer who selects videos by apparent popularity is not thereby selecting for accurate information, and may even be drawn toward less reliable content.

Of note, and in contrast to some earlier appraisals (including a study of mycetoma, another neglected tropical disease, in which longer videos were of higher quality [6], and an analysis of aspergillosis in which duration correlated strongly with all three quality scales [9]), we observed no association between video duration and quality. This suggests that, for leishmaniasis, length is not a useful heuristic for content quality, and that even brief videos from credible sources can convey reliable information while some longer videos do not. The strong inter-correlation among GQS, mDISCERN, and JAMA, together with good-to-excellent inter-rater agreement, supports the internal consistency of our assessments and the robustness of these conclusions.

These findings carry public-health relevance. Leishmaniasis disproportionately affects poor and rural populations in endemic regions, where accurate, accessible information is essential for prevention, early recognition, and care-seeking [1,2]. As digital platforms become more central to health information, the moderate quality and weak transparency of even professionally produced leishmaniasis content highlight a need for experts and institutions not only to create such content but to do so to higher standards, namely clearly stating authorship and credentials, citing reliable sources, disclosing conflicts, and dating their material. Collaboration between health authorities and platforms to promote and signpost high-quality, transparent content could improve the information environment for this neglected disease.

This study has several limitations. First, our search was limited to the three most relevant English-language search terms and to the 150 most-viewed eligible videos; less popular or non-English content, including the substantial volume of non-English videos we excluded, was not assessed, and our findings may not generalise to those audiences. Second, YouTube metrics such as views and likes are dynamic, so our data reflect the platform only as of the search date (15 June 2026). Third, the assessment instruments, while validated and widely used, involve a degree of subjective judgement; we mitigated this through independent dual rating, consensus resolution, and reporting of inter-rater reliability. Finally, we did not perform a detailed content analysis of the diagnostic or therapeutic accuracy of individual videos, which remains an important avenue for future work. Future research could incorporate non-English content, apply additional understandability and actionability measures, and examine short-form formats such as YouTube Shorts.

## Conclusions

YouTube videos on leishmaniasis are of moderate quality and reliability but limited transparency, and are produced almost entirely by professional and institutional sources. Quality is not reflected in video popularity, length, or age. These findings underscore the need for experts and institutions to produce not only more, but more transparent and clearly sourced, educational content on this neglected tropical disease.

## Data Availability

All data underlying the findings are fully available without restriction. The complete dataset, including all video identifiers, screening decisions, and reviewer scores, is deposited in figshare and available via DOI: https://doi.org/10.6084/m9.figshare.32677776.

## Declarations

### Author contributions

Kaweesi Calvin Nantalaga: conceptualization, methodology, software, data curation, formal analysis, investigation, visualization, writing – original draft, writing – review & editing. Nantege Alice: investigation (independent video assessment), validation, writing – review & editing. Both authors approved the final manuscript.

### Data availability

All data underlying the findings are fully available without restriction. The complete dataset, including all video identifiers, screening decisions, and reviewer scores, is deposited in figshare: https://doi.org/10.6084/m9.figshare.32677776.

### Use of generative artificial intelligence

Generative AI tools were used to assist with the development of the data-retrieval and analysis code and with language editing. They were not used to generate, screen, or score the study data, nor to draft the intellectual or scientific content. All video screening, assessment, and interpretation were performed by the authors, who take full responsibility for the integrity and accuracy of the work.

